# Creating Pathways for Change to Increase Modern Contraceptive Uptake in Rural Indonesia: Protocol for a Feminist Qualitative Study

**DOI:** 10.1101/2025.11.24.25340919

**Authors:** Rut Rosina Riwu, Frederick Ho, Sharon Greenwood, Cindy M. Gray

## Abstract

**Background:** Family planning program has been globally shown to reduce maternal mortality by reducing both total and high-risk pregnancies. Despite the national implementation of this program since the 1970s, Indonesia still faces many challenges in achieving family planning goals. Low modern contraceptive prevalence rate (mCPR) remains a problem that impacts public health, population growth, economy, and welfare issues. It should be tackled, especially in rural areas, with multifactorial causes and diverse needs. Various programs have been developed globally to overcome this problem; however, each region has different characteristics and demands that should be understood.

**Objectives:** This study aims to develop a theory of change by understanding rural women’s needs and actively collaborating with multiple participant groups to increase modern contraceptive uptake. The theory of change will also be informed by the views of four distinct stakeholders who is responsible for providing contraceptive services (i.e. policy makers at regency and provincial level) in order to make informed recommendations.

**Methods:** This feminist qualitative study embedding participatory action research principles adapts the first three steps of the six essential steps for quality intervention development. The target location is West Sumba Regency, one of Indonesia’s 100 lowest mCPR regencies and located in East Nusa Tenggara, which has the highest total fertility rate in Indonesia. Consisting of two rounds of data collection, this study includes different participant groups (i.e. rural women and men, mothers-in-law, religious figures, cultural leaders, midwives, family planning educators, and policymakers) with different strategies. To ensure data saturation and trustworthiness, we aim to recruit up to 45 participants through purposeful sampling, selecting participants based on the criteria for each group. The data collection methods are focus groups and semi-structured interviews. We will analyze the data using reflexive thematic analysis.

**Results:** The theory of change development focuses on women’s voices and incorporates various perspectives from rural communities, including the service providers and policymakers. Ethics approval has been obtained by the College of Medical, Veterinary, and Life Sciences (MVLS) Research Ethics Committee, University of Glasgow, UK, and the Public Health Faculty, University of Nusa Cendana, Indonesia. We anticipate that we will complete all data collections and analysis by December 2026.

**Conclusions:** The ultimate goal of this study is to develop a theory of change to create a meaningful change in contraceptive services in rural areas. This study will contribute to encourage rural communities to collaborate and empower rural women to overcome their reproductive health problems. By understanding the diverse contexts and specific needs of the rural population, the results will be essential to transforming family planning programs. In doing so, it will significantly enhance women’s reproductive health while also addressing and reducing health inequalities in rural areas.

## Introduction

The family planning program, established nationwide in Indonesia since the 1970s, has benefited women and children in numerous ways. First and foremost, it is an essential strategy to reduce maternal and child mortality by preventing unintended pregnancy, especially among high-risk groups, such as women aged less than 18 and more than 35, those who have more than three children, and those who have another child within two years of their last full-term pregnancy. If all women had access to modern contraception, one in three deaths related to pregnancy and childbirth worldwide could be avoided.^1^ Due to contraceptive use, it is estimated that between 523,885 and 663,146 maternal deaths were averted from 1970 to 2017 (a 37.5% - 43.1% reduction). If the contraceptive prevalence rate (CPR) were to rise from 63% in 2017 to 70% in 2030, and unmet contraceptive need were to fall from 10% to 7%, an additional 34,621 to 37,186 maternal deaths would be prevented (18.9-20% reduction). Furthermore, if Indonesia could reach an ambitious target of 75% CPR and 5% unmet need in 2030, 51,971 to 54,536 mothers would be saved, a 28.4-29.4% reduction in maternal deaths.^2–4^

In addition, family planning program can influence other health outcomes and economic development. The risk of low-birth-weight babies can be reduced by providing effective birth spacing methods. If all reproductive-age couples can access modern methods of contraception, not only unwanted and high-risk pregnancies, but also unsafe abortions can be prevented. When reproductive-age couples can manage their fertility, family circumstances could also be improved by minimising stress on household resources, including food, income and time. In fact, modern contraceptive uptake can also increase economic development by reducing family size.^3,4^

However, Indonesia continues to experience significant population growth, ranking fourth globally. In reaction to this challenge, Indonesia has implemented health programs focusing on reproductive health promotion and education, strengthening access to modern contraceptive services, and supporting informed choice in family planning. Regardless of these excellent efforts, the prevalence of modern contraceptive use stands at 60.4% in 2023, falling short of the ambitious 75% target set for 2030.^5,6^

As a part of family planning programs, modern contraceptive methods, which are more effective than traditional ones, aim not only to enhance maternal and reproductive health but also to uphold human rights. One of the key messages of the Safe Motherhood is that all women should have access to contraception to avoid unintended pregnancies.^7,8^ Thus, every individual has the right to access and choose from the advances in technology that enable them to select the most appropriate and suitable contraceptive method for their needs.^9^ However, in Southeast Asia, Indonesia has the fourth-highest unmet family planning need rate (11%).^10^

Some parts of the eastern region of Indonesia report highest unmet contraceptive needs, with a gap of 11.71% between western and eastern Indonesia. East Nusa Tenggara, located in the eastern region, faced greater challenges in family planning programs. The area had a lower participation rate in family planning programs, standing at 41.5% in 2023. Compared to other provinces in the eastern region, the difference is 7.5%; however, it increased significantly to 19.21% compared to other provinces in the western region.^6^ Nearly 34% of reproductive-aged couples in East Nusa Tenggara lack access to modern contraceptive services.^11^ Additionally, the majority of couples in this province are not modern contraceptive acceptors. Figures from 2021 to 2022 estimate that around 40% of couples used modern contraception. Thus, this province has the highest total fertility rate (TFR) in Indonesia (2.70).^6^

A variety of factors, both directly and indirectly, influence modern contraceptive uptake in different settings. Rural areas tend to have more complex related problems than urban areas, with the rural population reporting more barriers to accessing family planning services.^12^ Therefore, modern contraceptive use in rural Indonesia remains low, even though family planning programs have been established for more than 50 years. Building on previous studies that have identified the problem and its causes, our scoping review has highlighted behavioral factors and social support,^13–20^ accessibility,^1,20–23^ religious and cultural beliefs,^24–28^ and health system challenges.^29–32^ Rural and less educated women and communities with low wealth status need additional support to improve their reproductive health.^33^

Moreover, East Nusa Tenggara, which has many rural areas, is a multicultural province. It has cultures related to reproductive health that can positively and negatively impact women’s reproductive health. One challenge in rural areas is access to health facilities due to long distances and geographical barriers.^1,21–23^ In addition, women tend to be afraid of medical contraception’s side effects and be more trusting of shamans to help them control their fertility.^34,35^ On top of that, the patriarchal culture, which is still firm in most places, often makes women victims of gender inequality. A unique custom of marriage preparation in this place is the cultural gift (*belis*, in traditional language, meaning dowry) that must be given to the bride’s family and is determined by the bride’s status.^36–40^ The counts are different in every tribe, and it is one of the reasons why women must obey their husbands and in-laws, including for contraceptive decisions, as a previous study in our scoping review showed that the amount of dowry affected the number of children expected by parents-in-law.^41^

To increase modern contraceptive services, numerous reproductive health programs have been implemented globally, especially in rural areas where unmet contraceptive needs are still high. For instance, a male engagement family planning intervention in rural India showed that the Counselling Husbands to Achieve Reproductive Health and Marital Equity (CHARM) intervention represents a promising approach for challenging the root causes of women’s unmet need for contraception. It not only improves family planning use but also alters the male gender role ideologies that reinforce male reproductive control. In addition, educational intervention for religious leaders implemented in rural Tanzania increased community knowledge, demand for, and ultimately, uptake of family planning. Moreover, Family Health=Family Wealth (FH=FW), a multilevel intervention implemented in semirural Uganda, also showed positive effects on contraception services, increasing high-efficacy contraceptive uptake among couples wanting to delay pregnancy.^42–44^ Nevertheless, East Nusa Tenggara has distinct characteristics that must be considered in developing and implementing theories for contraceptive programs.

Our preliminary interviews with family planning providers in East Nusa Tenggara showed that the province has made significant efforts to implement the national family planning program, including integrating family planning services into national health insurance and involving religious leaders. Nevertheless, it is hard to be a successful family planning program if we only focus on the target and do not give attention to the quality of care of contraceptive services. Only after we focus on the quality of care will we achieve the target of family planning programs.^45^ Thus, to achieve family planning targets while providing quality of care along the continuum, we need to use a program theory approach to articulate the causal mechanisms underlying two changes: changed behavior and changed contraceptive status.^46^

Despite factors influencing the uptake of modern contraceptives in Indonesia being highlighted by previous studies in our first scoping review, there still remains a significant gap in research focusing on how program theories can effectively tackle the barriers to contraceptive use in rural communities. Addressing this gap is crucial to improving access and promoting family planning in these areas.^47^ Additionally, our second scoping review on interventions to increase modern contraceptive uptake in rural areas of Southeast Asian low- and middle-income countries (LMICs) found that the vast majority of studies have focused on urban areas. Limited studies are focused on increasing modern contraceptive use in rural Indonesia. This leaves a critical gap in our understanding of the unique socio-cultural and health system challenges that rural communities encounter.^48^ If we do not address these disparities, family planning programs risk remaining ineffective in rural Indonesia, thereby limiting access to essential reproductive health services for those who need them most.^10,49,50^

Building on the findings of our scoping reviews, which identified facilitators, barriers, and interventions for modern contraceptive uptake in rural areas of Southeast Asian LMICs, this study adopts a structured, theory-driven approach to intervention development. We use six steps in quality intervention development (6SQuID) to ensure a rigorous, contextually relevant framework for understanding the interplay among behavioral, socio-cultural, and health system factors. It offers a stepwise framework for designing an effective public health intervention, guiding the identification of causal mechanisms, defining targeted change pathways, and ensuring feasibility in real-world implementation.^51^

Moreover, to ensure the strategies developed are coherent, evidence-based, and aligned with the socio-cultural realities of rural Indonesia, the program theory approach provides a clear roadmap for linking intervention components to expected outcomes.^46^ By gathering insights from people with lived experience of modern contraceptive services, cognising their problems, and engaging with the community and related stakeholders, we can first gain a better understanding of the causal and contextual factors of modern contraceptive use across the full range of populations and stakeholders locally. We can clarify which factors are modifiable and have the most significant scope for change. Then, we can identify how to create a meaningful change in the modern contraceptive behaviors of rural communities.

Based on government data, local culture, our scoping review findings, and consultations with stakeholders, we are interested in developing a theory of change to improve family planning programs in rural areas of Indonesia by involving and empowering rural communities. This research directly supports the family planning program launched by the government, as the research-based theory of change will provide a clear, evidence-based framework for explaining how and why a specific intervention is expected to achieve intended outcomes. It will also help all involved actors contributing to family planning services achieve better results and create meaningful change by navigating the design, implementation, and evaluation of future interventions that are more effective and culturally appropriate. Overall, the theory of change will support well-designed interventions in the future that aim to increase modern contraceptive uptake in rural areas of Indonesia.

### Literature Review

The International Conference on Population and Development (ICPD) consensus on reproductive health has advocated for access to safe, effective, affordable, and acceptable contraceptive methods for all populations.^3^ However, the reality is that many reproductive-age women, especially those who are categorized as a hard-to-reach population, are confronted with significant challenges in accessing family planning services. A range of barriers, which have been well documented, such as geographical access, limited infrastructure and healthcare system, cultural norms, and lack of skilled providers, contribute to the low uptake of modern contraception. In addition, various strategies have been implemented and evaluated to enhance access to a variety of family planning services, especially in rural areas.^3,52^

Despite numerous studies exploring factors related to modern contraceptive use in rural areas of Southeast Asian LMICs, culturally appropriate and effective evidence-based interventions tailored to rural populations remain scarce and fragmented. To better understand existing evidence, two scoping reviews were conducted to examine facilitators, barriers and interventions related to modern contraceptive uptake in these settings. The findings highlight critical gaps that inform the need for this study.

Our first scoping review examined numerous barriers to modern contraceptive uptake in rural areas of Southeast Asian LMICs. Our scoping review meticulously selected 74 of the most relevant studies out of 13,442. Surprisingly, more than 65% of these studies are observational, underscoring the need for more in-depth qualitative research. Despite the potential of qualitative studies to provide profound insights into women’s contraceptive behavior,^53^ our review identified only 20 such studies, revealing a significant gap in this area of research, as many included studies recommended using a qualitative study design.

We adapted the socio-ecological model^54^ to describe the findings, as shown in **Figure 1**. Socio-cultural and religious barriers outweigh other factors to modern contraceptive services in rural areas. Even though women have access to modern contraceptive services, it is still arduous for them to decide and use the method as they need. Many considerations within families and society influenced women’s reproductive rights.

**Fig. 1.**
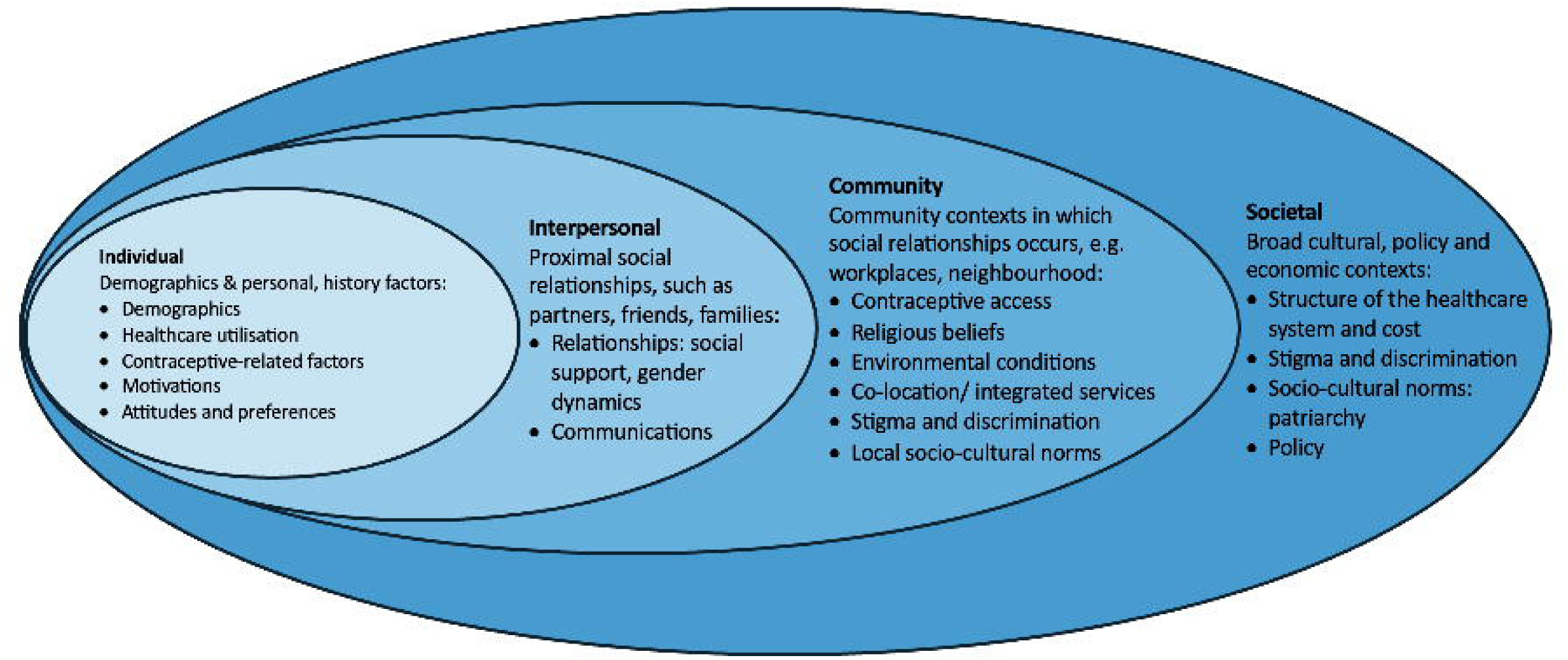
Identified factors related to modern contraceptive uptake based on socio-ecological model (SEM)

On top of that, our second scoping review, which examined interventions of modern contraceptive uptake in rural Southeast Asian LMICs, found that only six studies of 13,529 were included. Findings of the scoping reviews also reveal that while barriers to modern contraceptive uptake are well-documented, there is no evidence of effective, context-specific interventions for rural Indonesia, especially East Nusa Tenggara. Notably, although one study conducted an educational intervention that adopted religious and socio-cultural norms, none of the studies considered gender equality in their interventions. The outcome of most studies is women’s contraceptive practices, even though the intervention focused on educating men. Thus, this study will address these gender gaps to create a theory of change that could assist policymakers in developing a targeting strategy to increase modern contraceptive uptake in rural Indonesia.

### Aims

The overarching aim of this study is to explore modern contraceptive use in rural Indonesia with a focus on producing a theory of change to increase the uptake. To achieve this, we will identify: 1) the reasons why rural couples do not want to use modern contraceptive methods, 2) the main facilitators of and barriers to modern contraception uptake in rural areas and how they influence women’s contraceptive decisions, 3) change mechanisms that could be more effective and culturally appropriate to increase modern contraceptive uptake in rural areas based on the needs of rural populations; and will develop a theory of change to bring about the change at different levels and could maximize efficacy, acceptability, and impact.

## Methods

### Theoretical Frameworks

The socio-ecological model (SEM)^54^ focuses on mechanisms of change for individuals, families, groups, and communities, as well as the interplay among all those actors. Additionally, the Rainbow Model^55^ is a framework for assessing health inequalities that maps the interactions between the individual and the various layers of influences surrounding them. These theories are chosen for this study because people, in this case, rural couples, are part of systems. Their behavior is influenced by the environments in which people function: socio-cultural, physical, and other external environmental variables. On the other hand, individuals also influenced their environments, including the people around them. Furthermore, this model consists of several different levels, which are microsystem, mesosystem, exosystem, and macrosystem and can be conceived as a set of nested structures explaining how families, groups, communities and broader society, including other external environments or health system, contribute to rural couples’ contraceptive behaviors in a complex and interacting way.^46^

Empowerment emphasizes community participation and action research. It seeks to link individual strengths and competencies, natural helping systems, and proactive behaviors to social policy and social change.^56^ This is a process by which people exercize autonomy over their lives and can influence others who affect their decision-making. It emphasizes democratic participation, improvement, and self-determination. Considering one of the most potent driving forces in this framework —the sense of being part of something bigger —this theory is also chosen because it empowers participants to overcome their own problems, namely the low uptake of modern contraceptive methods. This theoretical framework will help us encourage people to adopt healthier behavior, which is accessing modern contraceptive services and practising their use by participating in decision-making and problem-solving. It will also help us to advocate for reproductive healthcare policies related to modern contraceptive services in rural areas.^46^

While SEM and the Rainbow Model will be used to address all factors contributing to rural couples’ modern contraceptive behaviors, empowerment theory will ensure a participatory research process and sustainable results.

### Study Design

Findings from our scoping reviews indicate that qualitative methods are needed to explore cultural and religious norms, social stigma and discrimination, and health system factors that influence modern contraceptive uptake in rural areas. Thus, this study is a feminist qualitative study embedding Participatory Action Research (PAR) principles that adapts the first three steps of 6SQuID,^51^ which are:

1. Define and understand the problem and its causes;
2. Clarify which causal or contextual factors are modifiable and have the greatest scope for change and who would benefit most; and
3. Identify the mechanisms of change.

After completing the first of 6SQuID’s steps in our scoping reviews, we proceed to the second and third steps in this study. We expect to gain a clear understanding of what the future intervention might look like and of how the developed theory of change will bring about change at different levels. Embedded in the principles of PAR, this feminist qualitative study is a form of research that meaningfully includes participants in every step of the research process. Unlike other forms of research, where researchers distance themselves from participants, this study involves participants, with the philosophy that participants are experts in their own lives, hold knowledge, and are able to collaborate.^57–59^

Focusing on partnerships between researchers and research participants in a collaborative effort, this study aims to overcome a specific problem: the low use of modern contraceptive methods in rural areas of Indonesia. Aligning with the principles of PAR, this study firstly encourages the active participation of the community in resolving the low uptake of modern contraceptive use. In addition, it is more focused on joint learning to uncover the causes of a phenomenon or problem rather than on scientific proof. Three main components of PAR, which are participation, action and research, go together in the following way: active and democratic participation of the people being observed and related stakeholders in the production of new knowledge through rigorous research that results in concrete action intended to create a meaningful social change within the community.^60–64^

### Study Setting

We chose West Sumba Regency, East Nusa Tenggara, Indonesia, for this project due to the fact that this place has a low mCPR (50%) and is among the 100 Indonesian regencies with the lowest modern contraceptive prevalence.^65^ It could represent the overall characteristics of the population needed, such as the balance between acceptors and non-acceptors. In addition, its location is far from the provincial capital. Located on a different island from the capital city and at the distant southeastern edge of Indonesia, most of Sumba Island is considered a rural area according to the World Bank’s rural criteria.^66,67^ On top of that, the strong culture of the community is regarded as a factor that might influence couples’ contraceptive behaviors.

Regarding the strong culture, Sumba is one of the areas in the world that still has long-standing practices of traditional slavery.^67^ Social hierarchy in Sumba culture consists of *Ratu* (the priest), *Maramba* (noblemen), *Kabihu* (free men/ordinary people/commoners), and *Ata* (enslaved people).^68^ Individuals of the noble caste are organized into various clans across Sumba Island. Being positioned at the bottom of the social hierarchy within the community, enslaved people are viewed as the property of nobles. They are obligated to serve their noble masters in myriad ways, including handling daily household chores, assisting with farming, and contributing to the production of hand-woven cloth.^67^ In fact, young, enslaved women often were a part of the dowry in ceremonial contexts, accompanying the bride to her new home with the groom.^36,69–71^

Moreover, Sumbanese also have a strong belief in *Marapu*, a local religion in the community that holds that ancestors’ spirits are gods. *Marapu’s* cultural identity distinguishes Sumbanese people and their traditions from those of other communities in Sumba island and other regions of Indonesia. It also underscores the uniqueness of Sumbanese cultural identity, expressed through cultural materials and various cultural activities (*adat* or tradition), such as the dowry (*belis*) tradition.^72^

In Marapu-derived traditions, the *belis* is a marker of respect for the bride’s family, prestige, and the social status of the bride and groom’s family. The male family gives animals (such as horses and buffaloes) and *mamuli* (gold or silver jewellery that resembles a woman’s womb, symbolising femininity and fertility and meant to honour women’s position). Meanwhile, female families give items such as weavings for men, sarongs and traditional women’s clothing. The number of items given varies according to social status and agreement, with the balance between them taken into account. This means the dowry system serves a positive purpose. However, this tradition raises several issues because it is considered a culture that makes women “chattel” so that they can be treated arbitrarily, for example, gender-based violence.^70,73^ Additionally, just like other patriarchal communities, married women in rural Sumba must live with their husband’s family; thus, they are more likely to be influenced by mothers-in-law and other family members for family planning decision-making and have less freedom to access the services.^74^

Given the various cultural factors that may influence rural couples’ contraceptive practices, we plan to conduct our project in West Sumba and include diverse groups as participants, as explained below.

### Participants and eligibility criteria

There are two rounds of participants recruitment for this study. For the first round of data collection, this study will involve up to 40 participants (rural women as key informants, rural men, mothers-in-law, religious figures, cultural leaders, midwives, and family planning educators) and 10 local collaborators (co-participants). Additionally, for the second round, this study will involve up to five informants (policymakers in different level). We will select ten participants from all participant groups in the first round to serve as our local collaborators (co-participants). Therefore, given the theoretical saturation, our target is 40 to 45 informants, as shown in Figure 2.

**Fig. 2.**
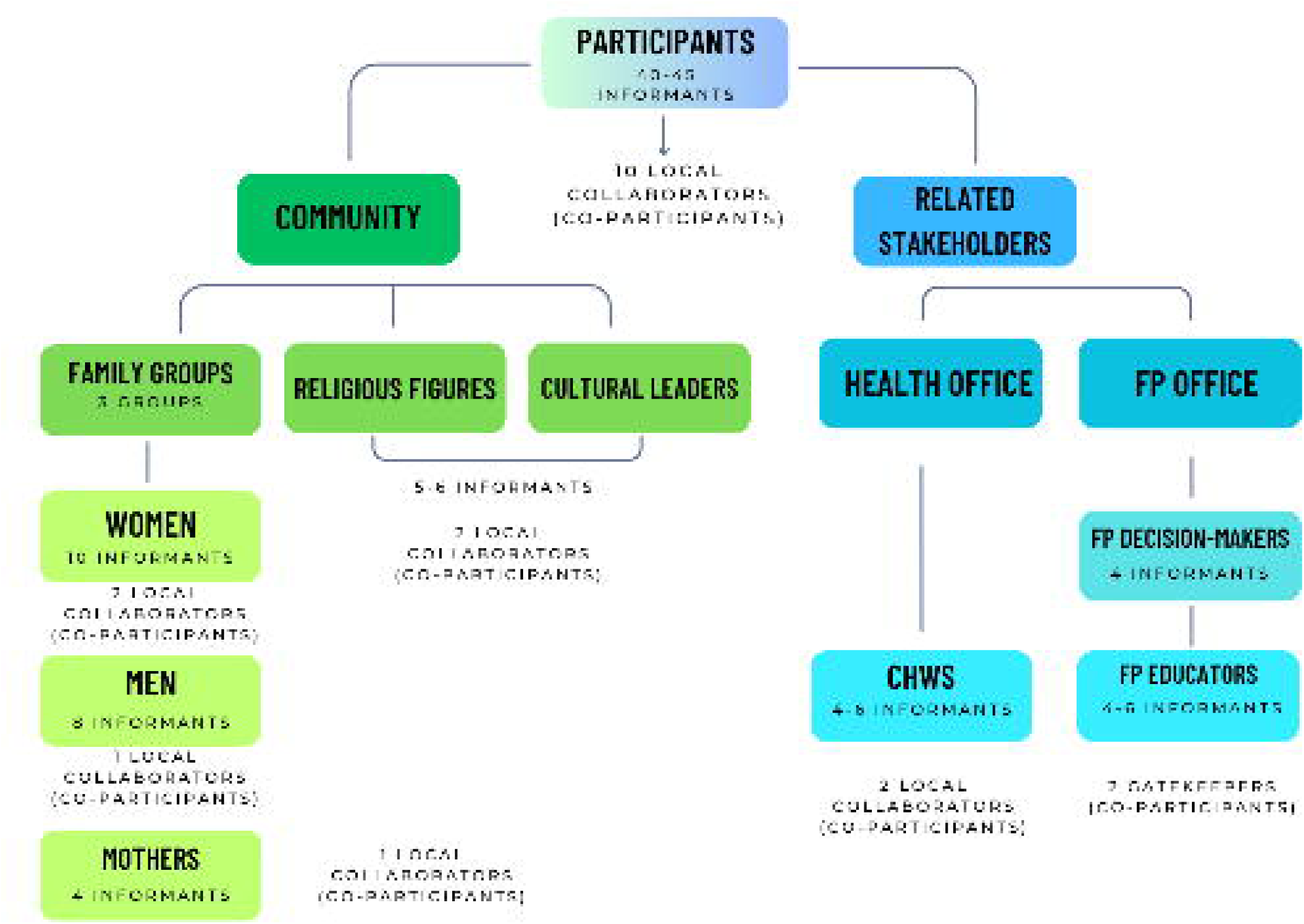
Research Participants

#### Women and Men

Women and men living in the study location who have the characteristics listed in **Table 1** are eligible to participate. Because this is feminist research, eligible rural women served as key informants. Given confidentiality concerns, the women and men participant groups did not come from similar households, villages, and districts. The data collection, focus groups, were held separately in different districts.

**Table 1.** Eligibility criteria for the participation of couples.

| Inclusion criteria | Exclusion criteria |
| --- | --- |
| • Reproductive age (15 – 49 years)<br>• Registered to the Population Control, Women's Empowerment and Child Protection Office of West Sumba Regency as couples of childbearing age who do not use modern contraception<br>• Those who do not use modern contraception, especially those who have a husband/partner or family who does not want them to use it<br>• Willing to participate in the study<br>• Permanent residents of the study location. | • Cognitive impairment (husband or wife)<br>• Those who do not use contraception because of particular conditions, such as infertility or live separately from their partner (do not use contraception objectively).<br>• Intend to move in the next 12 months, or either spouse refuses participation. |

#### Mothers

Eligible participants are mothers-in-law who live with married or unmarried couples in the same house in the study location and are permanent residents. Considering the crucial role of mothers in determining customary decisions,^36^ they are preferred participants in this study. If they do not have a cognitive impairment and would like to participate in this study, they will be included.

#### Community health workers (midwives)

Eligible midwives are those who 1) registered in the Health Office of West Sumba Regency, 2) have experience as midwives at a community health center or as village midwives, 3) have served the community for at least two years, and 4) are willing to participate in this study.

#### Family planning educators

Eligible family planning educators are 1) those who are registered in the Ministry of Population and Family Development; 2) have experience in promoting the community about family planning program and contraception in the target location for at least two years.

#### Religious leaders/figures

Eligible religious leaders/figures are 1) those who have experience as a leader/figure in their own religion or belief for at least five years; 2) those who are trusted by the congregation in leading religious practices in their place of worship and giving spiritual guidance. Considering the need for diverse representation, participants from this group will come from different religions or beliefs, including local or traditional beliefs, in the target location. Those who are willing to participate in this study will be included.

#### Cultural leaders

Eligibility criteria for participation as cultural/community leaders are 1) those who the community has trusted for years in preserving cultural practices and traditions, addressing cultural needs of the community, and fostering social relationships within the community; 2) those who live in the study location as permanent residents.

#### Policymakers

These participants are those who served as policymakers in health and family planning programs in West Sumba Regency and East Nusa Tenggara Province.

The sampling technique for this feminist qualitative research is purposeful sampling, considering all informants must come from these groups: (a) childbearing age women not using modern contraception, (b) men not using modern contraception, (c) mothers-in-law, and (d) community health workers (midwives), (e) religious leaders/figures, (f) cultural leaders, (g) family planning educators, and (h) family planning program decision-makers in the regency and province level as listed in **Figure 2**. Three selection criteria are (a) information-rich informants, (b) do not have cognitive impairment and (c) willingness to participate in the research process.

### Protecting Privacy and Confidentiality

Protecting privacy and confidentiality is essential to ensuring women’s safety and data quality, especially for sensitive research that studies private lives, deeply personal experiences, and sacred things.^75,76^ Given that the participants are the indigenous population of West Sumba, who are vulnerable and difficult to reach, we carefully ensured confidentiality and used different strategies throughout the study. Firstly, by separating each family group (women, men, and mothers-in-law). Each family subgroup did not come from a similar village. The women and mothers group came from a different district than the men’s group. Each focus group included participants from different villages. Considering the cultural conditions and risks that women participating in this research might face, we separated family groups to different locations not only during data collection but also during participant selection. By doing so, the possibility of being known by other family groups and their families could be reduced.^77–79^

In addition, the family groups are not drawn from the same household or house to reduce participants’ discomfort when discussing sensitive topics in focus groups and to address participants’ fears that their spouse or mother-in-law might learn the nature of the discussion between research staff and participants. Furthermore, focus groups for family groups were held in different places and at different times. Data collection on women, men, and parents was conducted separately in a safe, neutral location, depending on participants’ willingness, at different times, with discussion with the family planning educators serving as our gatekeepers. Not only family groups but also all participant groups were involved in selecting the focus group location. Moreover, anonymity or different names will be used to remove personal identifiers and maintain the confidentiality of personal data.^78,80^

It is ensured that participation in this study poses no risks to participants. Given confidentiality concerns, participants, especially local collaborators and their organisations, will not be named in subsequent write-ups or materials submitted for publication. They will not be contacted in the following write-ups and materials submitted for publication. However, in line with participatory action research principles, it can be difficult to ensure confidentiality when participants or stakeholders are willing to be mentioned in the results. Therefore, the final decision depends on the participants’ decision to be named in the results.

### Data collection

Data collection for the first stage is planned for August – September 2025 and the second stage is planned by December 2026 as showed in **Figure 3**.

**Fig. 3.**
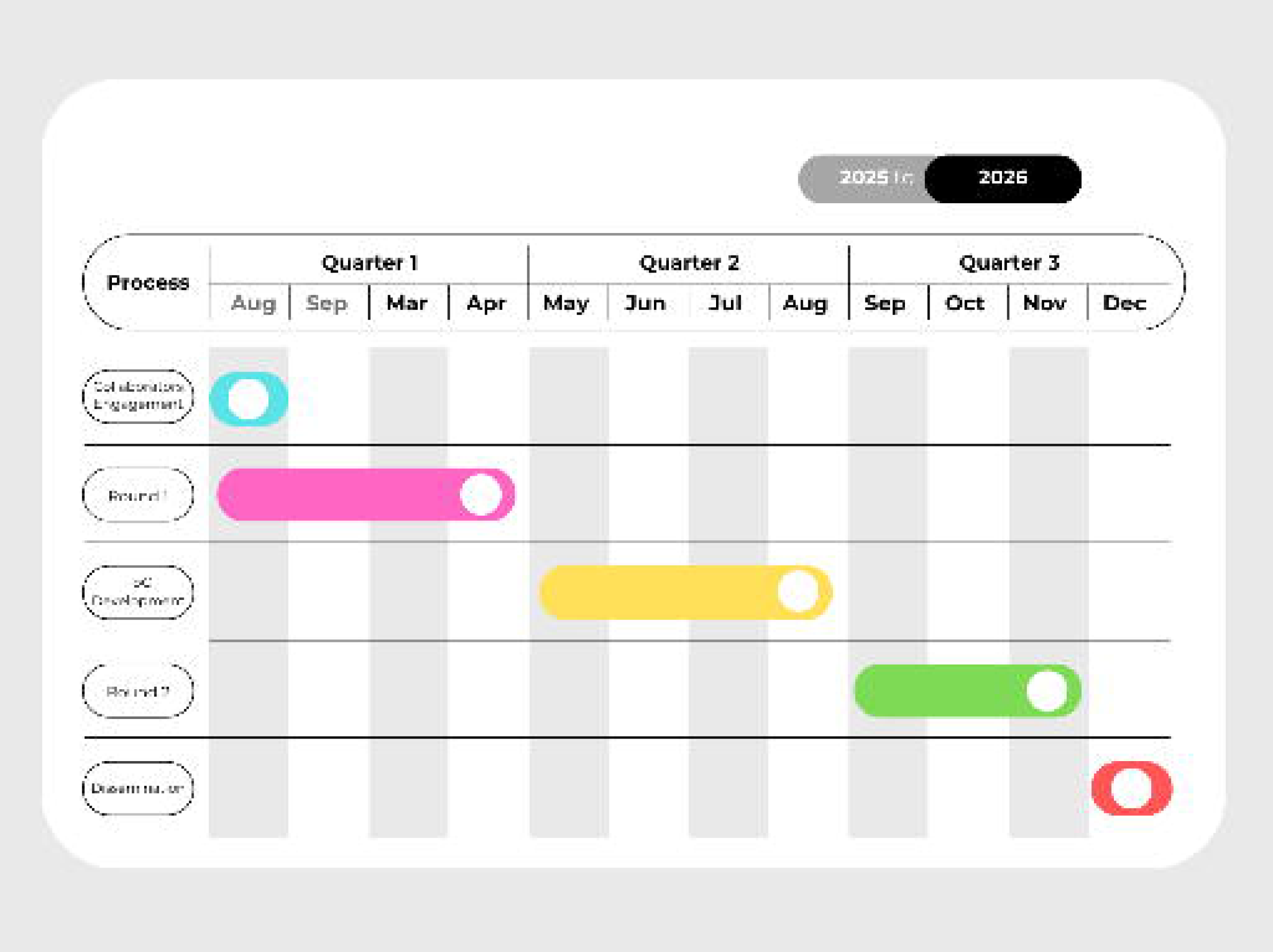
Project Timeline

Data collection methods are focus groups and semi-structured interviews. Focus groups are particularly effective at eliciting data on a group’s cultural norms and, importantly, at generating broad overviews of issues of concern to the cultural groups or subgroups represented. In addition, focus groups are effective for capturing a range of perspectives within that community or subgroup. This qualitative data collection is often used to determine the kind of service a particular population would like. Because focus groups seek to illuminate group opinion, this method is exceptionally well-suited for sociobehavioral research used to develop and measure services that meet the needs of a given population.^80,81^ For these reasons, we will use focus groups to gather data from all participant groups except the family planning policymakers. Semi-structured interviews will be used to collect data from family planning policymakers.

Before data collection with women and mothers, a vignette, a real-life scenario or story about a maternal health problem related to not using modern contraceptive methods, was presented to them. This vignette is a video of “*Why did Mrs X die? Retold*”, which has been remade by Professor Mahmoud Fathalla.^82^ This vignette aims to elicit participants’ responses, attitudes, and perceptions. Considering the complex and sensitive topics of this research, a vignette is used as a tool to help us explore in a direct, non-threatening way, allowing participants to respond to the given scenario before sharing their personal experiences. Our preliminary interview with rural couples from the study area revealed a common assumption that not using modern contraception had not been considered a problem. This vignette is a crucial tool to help participants understand that not using effective contraceptive methods is a significant health issue, potentially leading to severe problems like maternal deaths.

To ensure data collection is accurate and trustworthy, we have additional members as mentioned below.

#### Gatekeepers

Considering the principles of PAR, immersion in the community is a crucial step before beginning data collection and throughout the study. To immerse ourselves in the community, we have two gatekeepers who served as family planning educators in the study area. These gatekeepers, deeply connected to the community, will help facilitate access to the research site and participants, build trust and mediate relationships to ensure engagement with the target population or setting. One has been helped since the preparation step, while the other will help select participants. As part of the community that has worked to educate people about modern contraception, they will help us ensure the study is successful.

#### Research assistant and translator

Additionally, the support of a research assistant and two translators, who are native to and reside in the study location, will be crucial in our data collection efforts. Their role is to support various aspects of the research, from data collection to dissemination, and help us immerse ourselves in the community, a crucial aspect of our research, especially translators who facilitate communication between researchers and participants, ensuring that it is not just clear and contextually accurate, but also culturally appropriate, thereby enhancing the quality and depth of our research.

#### Psychologist

Furthermore, to ensure participants’ well-being and the integrity of the research, we have one psychologist who will help us in several ways. First, supporting participants’ well-being emotionally when discussing sensitive topics; second, providing immediate psychological support or referrals if participants experience distress during or after the research; third, giving sensitive training for team members before data collection for identifying distress; fourth, facilitating data collection by co-moderating discussions and observing emotional cues; interpreting emotional responses and importantly, addressing bias in data analysis to maintain the research’s integrity.

### Research stages and procedures

The steps of PAR are to develop a co-generative dialogue with the community of interest in immersion and problem identification, decide on a research strategy and process, and use joint, critical analysis to enact change. In conjunction with these steps and the principles of feminist research methodology, this study comprises several stages, as showed in **Figure 4** and explained below.

**Fig. 4.**
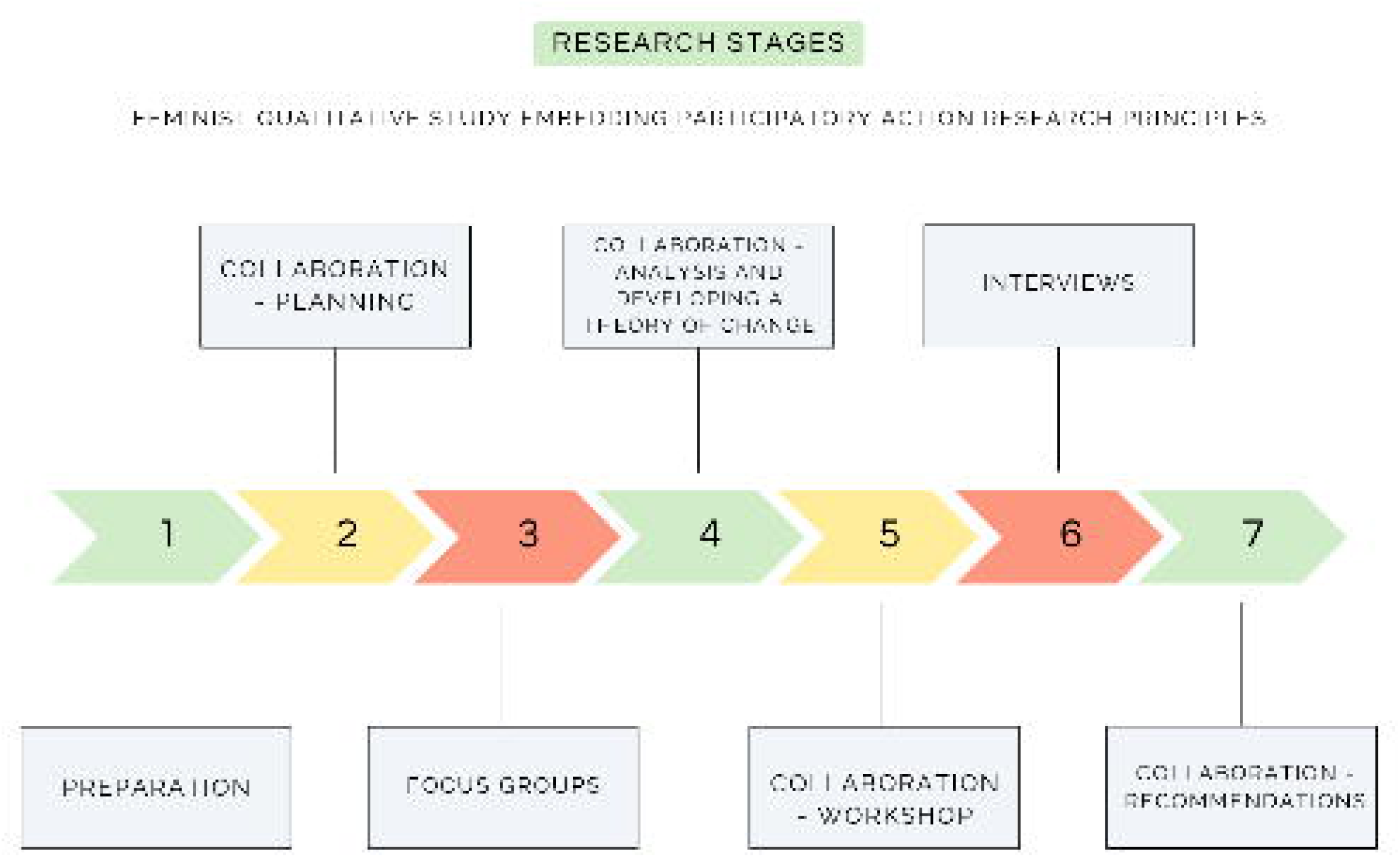
Research Stages

#### Preparation

a. Determining research topic, issues, goals, methods, setting, and target community.
b. Consulting these with the family planning chief and staff at the province level (National Population and Family Planning Board of East Nusa Tenggara) and regency level (West Sumba Population and Family Planning Board of West Sumba)
c. Writing: problem analysis, research proposal, protocol, and scoping reviews.
d. Applying for ethical clearance.
e. Ethical and sensitivity training for field work team members before data collection.

#### Collaborators Engagement

a. Identifying and recruiting local collaborators from the target community and relevant stakeholders, that are the Office of Population Control, Women’s Empowerment and Child Protection (DP5A) of West Sumba Regency; the Health Office of West Sumba Regency; and the Ministry of Population and Family Development/National Population and FP Board Representative of East Nusa Tenggara Province. Identification of local collaborators began with the local family planning educator group (under the Office of Population Control, Women’s Empowerment and Child Protection (DP5A) of West Sumba Regency). At the end of this step, local collaborators will be representatives from all participant groups.
b. Transferring knowledge, research topic, problem analysis, research questions, goals, and methods to gain trust and support.

In this step, we went to the Population Control, Women’s Empowerment and Child Protection Office and invited them to collaborate. In this first initial meeting, we shared the research topics, research questions, goals, and methods. Then we discussed with all family planning educators to select the target location and potential participants. Based on our discussion, two districts were selected, each located more than 30 km from the capital, and two family-planning educators are involved as our gatekeepers. We shared information sheets with our gatekeepers, who reshared them with potential participants. Our gatekeepers helped us select two midwives from different districts as our study locations, who then helped us select participants. After collaboratively selecting the participants with gatekeepers and midwives, we began the first round of data collection in August 2025.

#### Data Collection Round 1

This first round of data collection aims to identify which causal or contextual factors and acceptable mechanisms are modifiable, have the greatest scope for change, and who would benefit the most (the second step of 6SQuID). We will identify barriers to modern contraceptive uptake and acceptable mechanisms of change.

The methods are focus groups with women of childbearing age (n = 8 – 12; two focus groups), men (n = 8 – 12; two focus groups), mothers-in-law who live with reproductive-aged couples (n = 4 – 6, one focus group), midwives (n = 6 – 8, two focus groups, each involves midwives who serve in different district), religious figures and cultural leaders (n = 4 – 6, one focus group), and family planning educators (n = 4 – 6, one focus group), and at least one interview with traditional religious leaders (*Rato* in traditional language).

#### Data Analysis of Round 1

a. Familiarising with data, coding, categorising into themes of key causal or contextual factors influencing modern contraception use of rural couples in West Sumba, and acceptable mechanisms of change that could increase contraceptive behavior positively, then theorising.
b. Triangulating and gaining feedback from the family planning providers at the regency level.
c. Involving local collaborators in the analysis process by member checking and sharing the findings to gain their feedback.

#### Developing a theory of change

This stage aims to decide on the mechanisms of change. In this stage, we will develop a theory of change based on the findings from the first round to understand how to create a meaningful change to increase modern contraceptive behavior in different levels. This theory of change is expected to be a guide in future interventions to increase modern contraceptive uptake in the study location, and probably other rural areas.

#### Workshop

After developing the theory of change, we plan to undertake a community workshop that aims to share the developed theory of change with all participants, including local collaborators, and to give them a voice, allowing them to share their thoughts and opinions on the change mechanisms outlined in the theory of change. Their insights and perspectives are invaluable in shaping the direction of this research.

#### Data Collection Round 2

After conducting the workshop, we will continue the second round of data collection to determine the mechanisms of change and clarify how to deliver them. This second round is designed to gather information from family planning decision-makers about their thoughts and opinions on the developed theory of change and what needs to be done to bring about the change, in the form of semi-structured interviews (n = 4 – 5). Based on the results of previous rounds, this round can also reflect what can be adopted and what can be changed.

#### Data Analysis of Round 2

a. Familiarising ourselves with data, coding, categorising into themes of mechanisms of change and the implementation, and theorising.
b. Involving local collaborators in the analysis process by sharing the results in a community meeting.
c. Triangulating and gaining feedback from the regency and province-level family planning policymakers.

#### Dissemination and Advocacy

a. Writing reports and recommendations
b. Presenting the results to the family planning policymakers at the regency and province levels.
c. Advocating the developed theory of change to increase modern contraceptive uptake in rural areas.

### Data analysis

The focus groups and interviews will be transcribed verbatim, and reflexive thematic analysis will be used to analyze the data by identifying themes and patterns of meaning across datasets in relation to the research questions, and emphasising the role of researcher self-awareness and critical reflection throughout the analysis process.^83^ The analysis process consists of several steps: familiarising ourselves with data, generating initial codes, searching for themes, reviewing themes, defining and naming themes, and theorising and producing the report.^53,84^

To support the analysis, we will use NVivo, a qualitative data analysis software, to structure and store the data. This software will facilitate the identification of codes, the creation of analysis themes, and the initial exploration of all written documents arising from the focus groups and interviews.

### Data management

All focus groups and interviews will be recorded in audio. They will be transcribed, analyzed, and used to develop the theory of change. The study findings will also be published in an open-access journal. Data, including personal and special category data, will be stored securely under a password and used for research purposes only. Data-sharing agreements have been secured for external team members, including the transcriber. Given the qualitative nature and the potential for identifiable and sensitive information, sharing raw transcripts is not appropriate. However, de-identified data supporting published findings will be available after study completion in accordance with the University of Glasgow’s ethical approvals and data-sharing policies.

All data will be managed in accordance with the University of Glasgow’s Code of Good Practice in Research and Intellectual Property Policy. The University retains the right and responsibility to ensure secure storage of valuable data. Data must comply with institutional and ethical requirements. In line with University policy, all research data will be retained for ten years after project completion.

### Rigour

The following strategies will be used to ensure the project’s internal validity, credibility, and accuracy: first, several forms of triangulation of data collection techniques from interviewers and analysts; second, audit strategies, for instance, meaningful member checking or feedback from participants and feedback from critical experts; and third, transparent and thorough descriptions of the research process and methodological decision-making.^85,86^

### Ethical considerations

The College of Medical, Veterinary, and Life Sciences (MVLS) Research Ethics Committee of the University of Glasgow, United Kingdom, issued ethical approval in August 2025 (200240450), and the Public Health Faculty of the University of Nusa Cendana, Indonesia, issued ethical approval in May 2025 (001922/KEPK FKM UNDANA/2025).

All participation in focus groups, interviews, and collaboration is voluntary. All participants will receive an information sheet and a verbal explanation of the study. Written informed consent is obtained prior to the commencement of the focus groups and interviews. For participants with low literacy, the consent form is read aloud, and a thumbprint is accepted in place of a signature, witnessed by a person appointed by the participant. Participants’ consent is verified orally at the end of the session, and they are reminded to contact the research team lead to withdraw any or all of what they have said after the focus group or interview. Consent forms are stored securely and separately from data.

At the end of the first stage of data collection, we continue recruiting co-participants (local collaborators). A written and signed working agreement is provided to 10 participants who are willing to serve as local collaborators after discussion.

### Dissemination plan

At the completion of the study, we will present the findings, especially the theory of change and future action plans, to relevant stakeholders advocating for equitable reproductive health actions and policies, published in peer-reviewed journals, and presented at international conferences. Findings will be presented in a summarized form without any identifying information.

## Results

This study was funded in 2024 as part of the first author’s doctoral study and enrollment has not been completed yet since this study consists of two rounds of data collection. Currently, we are recruiting local collaborators to co-develop the theory of change based on the findings from the first round of data collection that was completed in September 2025. Data analysis for the first round is currently underway, and the results are expected to be disseminated to the related stakeholders in 2026. We anticipate that we will complete all rounds of data collection and analysis by December 2026 and submit the results for publication in 2027.

## Discussion

### Principal Findings

This study focuses on the needs of rural women in developing a theory of change to create a meaningful change in modern contraceptive services in Indonesian rural areas. Ethical approval has been obtained by the College of Medical, Veterinary, and Life Sciences (MVLS) Research Ethics Committee of the University of Glasgow, United Kingdom and the Public Health Faculty of the University of Nusa Cendana, Indonesia. Collaborating with rural communities in the research process is built on a philosophy that participants hold knowledge and can be involved.^57–59^ This study is crucial because it empowers rural communities to take charge of their reproductive health, make decisions and solve problems related to accessing modern contraceptive services. Furthermore, the theory of change will guide stakeholders in designing more effective, culturally appropriate family planning programs that foster empowerment among rural populations.

The expected outputs of this study include: 1) identification of the facilitators of and barriers to modern contraceptive uptake of rural women and men in Indonesia by understanding their perspectives; 2) identification of the causal or contextual factors and acceptable mechanisms to increase modern contraceptive services in rural areas of Indonesia based on rural women’s and men’s voices; 3) identification of other factors related to modern contraceptive uptake based on other perspectives apart from rural women and men, which are parents/in-laws, religious leaders, cultural/community leaders, and community health workers, including village midwives and family planning educators that can influence the theory of change and its impacts on rural women and men; 4) identification of modifiable causal or contextual factors with the most significant scope for change and who would benefit the most; 5) development of a theory of change based on the voices rural women that exclusively aims to empower them to adopt a positive modern contraceptive practices; and 6) determination of the mechanisms of change based on the theory of change and the way to deliver them at different levels by understanding related stakeholders’ perspectives.

### Conclusions

The ultimate goal of this study is to develop a theory of change based on collaboration with rural community to give a positive impact on family planning programs in rural Indonesia by increasing reproductive health knowledge, women’s empowerment and men’s involvement. By doing so, we anticipate that access to and use of modern contraceptive services will increase, whilst unintended pregnancy and maternal mortality will decrease.

While this study provides valuable insights into family planning programs, the major anticipated limitation is that the findings might not be generalisable to other rural communities with different cultural conditions. Nevertheless, conducting this study and disseminating its findings may help develop future interventions based on program theory, which provides a logical, evidence-based foundation.^46^

## Data Availability

Datasets have not been fully generated or analysed during the current study. Deidentified research data will be made available under the University of Glasgow, following the University’s Code of Good Practice and ethical standards, upon completion of the study and publication.

## Acknowledgments

We are grateful to the assistance of the Ministry of Population and Family Development/National Population and Family Planning Board for East Nusa Tenggara Province, Indonesia and the Population Control, Women’s Empowerment and Child Protection Office of West Sumba, Indonesia. We also thank Marvelous Ruly Pulung Tana, M.Kes. for his assistance with the data provision and the recruitment phase.

We used the generative AI (artificial intelligence) tool Grammarly by Grammarly Inc.* to suggest language improvements within the manuscript, Chat GPT by Open AI** to find and identify articles that are likely to be related to the research reported in the manuscript, and DeepL*** for translation.

* Grammarly. (2025). Grammarly [Large language model]. https://app.grammarly.com

** OpenAI. (2025). ChatGPT [Large language model]. https://chatgpt.com

*** DeepL. (2025). DeepL [Large language model]. https://www.deepl.com/en/translator

## Funding

This research protocol is part of the doctoral thesis of the first author under the supervision of the co-authors, Dr Frederick Ho, Dr Sharon Greenwood, and Prof. Cindy Gray of the University of Glasgow. The Indonesia Endowment Funds for Education (LPDP) supported this study, and the funder had no involvement in the study design, data collection, analysis, interpretation, or the writing of the manuscript.

## Author Contributions

All authors read and approved the final manuscript. All authors also contributed to the conceptualisation and methodology. First author contributed to project administrations, resources and writing original draft and editing. All co-authors contributed to supervision, review, and editing the manuscript.

## Conflict of Interests

The authors declare that they have no competing interests.

## Multimedia Appendix

Topic guide and questioning route of focus group and interview

## Abbreviations

6SQuID: Six steps in quality intervention development
CPR: Contraceptive prevalence rate
mCPR: Modern contraceptive prevalence rate

## Notes

### Competing Interest Statement

The authors have declared no competing interest.

### Funding Statement

This study is funded by the Indonesia Endowment Funds for Education (LPDP) and is part of the doctoral thesis of the first author under the supervision of the co-authors. The funder had no involvement in the study design, data collection, analysis, interpretation, or the writing of the manuscript.

### Author Declarations

Ethics committee of the College of Medical, Veterinary, and Life Sciences (MVLS) Research Ethics Committee, University of Glasgow, UK, and the Public Health Faculty, University of Nusa Cendana, Indonesia gave approval for this work.

### Summary of Updates

Section on Methods updated to clarify, and supplemental files updated.

